# Smoking, e-cigarettes and the effect on respiratory symptoms among a population sample of youth: retrospective study

**DOI:** 10.1101/2022.03.10.22272186

**Authors:** M Chaiton, M Pienkowski, I Musani, SJ Bondy, JE Cohen, J Dubray, TE Eissenberg, P Kaufman, MB Stanbrook, R Schwartz

## Abstract

**Introduction:** E-cigarettes have been steadily increasing in popularity, both as cessation methods for smoking and for recreational and social reasons. This increase in vaping may pose cardiovascular and respiratory risks. We aimed to assess respiratory symptoms in youth users of e-cigarettes and cigarettes.

**Methods:** A cross-sectional survey design was utilized to assess Canadian youth aged 16-25 years old. Participants were recruited from the Ontario Tobacco Research Unit Youth and Young Adult Research Registration Panel November 2020 to March 2021. A total of 3,082 subjects completed the baseline survey. Of these, 2660 individuals who did not have asthma were included in the analysis. The exposure of interest was vaping dose, pack equivalent years, equivalent to cigarette pack years incorporating number of puffs per day, number of days vaped per month, and number of years vaped. Respiratory symptoms were measured using the five-item Canadian Lung Health Test. Poisson regression analyses were performed while adjusting for demographic confounders, stratified by smoking status. A non-stratified model tested the interaction of status and vaping dose and the effect of vaping device used was assessed among ever vapers. Analyses controlled for demographic characteristics, use of cannabis and alcohol, and survey date.

**Results:** Each additional puff year increased the rate ratio of respiratory symptoms by a factor of 11.36 (95%CI: 4.61-28.00; p<0.001) for never smokers, but among current daily smokers higher pack equivalent years were not associated with more respiratory symptoms (0.83; 95% CI: 0.23., 3.11). Among current vapers, those using pod-style devices were more likely to have more respiratory symptoms (1.25; 95% CI: 1.08, 1.45) after adjusting for dose.

**Conclusions:** Vaping is associated with an increased risk of reporting respiratory symptoms among never smoking youth and non-daily ever cigarette smokers. Use of e-cigarettes among non-smokers should be discouraged.

**Funding:** Canadian Institutes of Health Research

## Introduction

Cigarette smoking has long been known to be associated with poor respiratory health outcomes, including lung cancer, chronic obstructive pulmonary disease (COPD), and severe asthma.^1^ In the past several decades, the prevalence of cigarette smoking has decreased drastically across North America. Even so, it remains prevalent with an estimated 14.8% of Canadians smoking cigarettes in 2019, which is slightly higher than rates of 14.1% in the United Kingdom and 14.0% in the United States.^2,3,4^

While the use of cigarettes has become less common, electronic cigarettes (e-cigarettes) have quickly gained global popularity as both smoking cessation aids as well as recreational nicotine delivery devices.^5^ This rise in use has been especially prominent amongst youth, with several studies observing large increases in the prevalence of youth e-cigarette use in both the United States and Canada within the past decade, with 15% of youth in Canada aged 15 to 19 reporting past 30-day use of e-cigarettes in 2019, compared to only 6% in 2017.^6,7,8^ Despite being initially introduced as a supposedly safer alternative to cigarettes, e-cigarettes have been found to be associated with some health-related harms similar to cigarettes.^9,10^ Further, the use of e-cigarettes has been associated with additional harms unique to the electronic devices.^10^ Literature surrounding the health impacts of e-cigarette use is still evolving, but existing research has identified both short- and long-term respiratory harms due to e-cigarette use.^11^ However, existing research primarily includes adult samples, limiting generalizability to youth populations, which differ biologically, psychologically, and sociologically. Further research is thus necessary to assess the effect of vaping frequency on risk of respiratory symptoms, especially among youth who are at highest risk of vaping initiation. Additionally, the association between vaping device type/e-cigarette liquid flavour and respiratory symptoms must be assessed as different devices provide varying doses of vapor per puff, which may impact risk of respiratory symptoms among users of certain device types, even with a lower vaping frequency.

In addition, many individuals engage in dual use of cigarettes and e-cigarettes.^12,13^ Motivations for dual use vary, with research suggesting that individuals who initiate the use of e-cigarettes with the goal of smoking cessation are more likely to engage in dual use than to engage in e-cigarette use alone.^12^ This may be the case because smokers who use e-cigarettes as smoking cessation aids may not fully transition from smoking to vaping, despite often beginning with the intention to quit smoking entirely.^12^

Research surrounding the health impacts of the dual use of cigarettes and e-cigarettes remains limited. Results from existing studies have been mixed, with some finding that dual use leads to mitigated health-related harms due to the overall decrease in the use of cigarettes.^12,13^ Other research has found that the potential mitigation of health-related harms as a result of switching from cigarettes to e-cigarettes are negated in those who continue to smoke cigarettes alongside using e-cigarettes. This may be observed as no difference in health-related harms between dual users and single product users or increased harms among dual users. Notably, most of these studies focus on the health-related impacts of either smoking or vaping, but not both.^14-18^ Finally, much research remains inconclusive surrounding the harms associated with the dual use of cigarettes and e-cigarettes, as well as with regards to how these impacts differ between individuals who are solely users of either cigarettes or e-cigarettes.^12^

The current analysis sought to use survey data to understand better the association between e-cigarette use and self-reported respiratory symptoms among youth, along with the association between self-reported respiratory symptoms and e-cig dose (number of puffs per month), vaping device, e-cigarette liquid flavour, and years vaped. In doing so, we seek to add to a growing body of research regarding the extent of harms related to the use of e-cigarettes. Additionally, the study aims to assess the interaction associated with the dual use of cigarettes and e-cigarettes on respiratory symptoms. Based on current research on both vaping and smoking cigarettes, we hypothesized that individuals who smoked cigarettes regularly and those who vaped regularly would report more respiratory symptoms compared to non-smokers and non-vapers. We further hypothesized that dual use of cigarettes and e-cigarettes would be associated with more respiratory symptoms compared to vaping alone, but lower compared to cigarette smoking alone, due to satiation of cravings that would decrease cigarettes smoked daily. However, this assumption of decreased cigarette intake may be incorrect, as some studies reported lower levels of smoking in dual users, while others reported no notable change in number of cigarettes smoked, though fewer respiratory symptoms were still observed.^18^ To further this understanding, we also considered potential confounding factors in the relationship between smoking and respiratory health, including age, sex, education level, and other substance use.

## Methods

### Data Source

Participants were recruited from the Ontario Tobacco Research Unit Youth and Young Adult Research Registration Panel November 2020 to March 2021 (Pienkowski, 2021). Panel recruitment was completed via social media advertisements from August 2020 to February 2021. Panel participants completed a recruitment survey and provided information for future contact. From the panel, adolescents and young adults aged 16-25 living in Canada were eligible for the survey. Targeted recruitment by email was used to balance the survey respondents for vaping status (never, ever (at least one puff), past 30 days), smoking status (never, ever, past 30 days) and age (16-18, 19-25). Participation rate was 60%. Younger females, those with higher education, and those who had never used e-cigarettes or cigarettes were more likely to participate. There were 3082 youth who completed the baseline survey including 396 individuals reporting a diagnosis of asthma. The University of Toronto Research Ethics Board provided ethics approval.

### Measures

The primary exposure variables was pack-equivalent years. This value was calculated equivalents to the pack-years calculation for cigarettes. The number of reported puffs per day on days vaped (divided by 10, the standard number of puffs in a cigarette) was multiplied by the number of days vaped per month (divided by 30 to provide an average daily use). This value was then divided by 20 to convert to packs (20 cigarettes per pack) and then multiplied by the number of years of vaping report (current age-age at first vape).

The outcome variable was the self-reported occurrence of adverse respiratory symptoms. Data on five respiratory symptoms were collected, with survey respondents identifying any symptoms they experienced in the past four months (yes/no). The five symptoms were coughing regularly, coughing up phlegm regularly, feeling out of breath from even simple chores, wheezing when exerting oneself (e.g., through exercise or going up the stairs), and getting many colds (specifically those that take longer to recover). The five questions were combined to form an overall respiratory symptoms variable, the Canadian Lung Health Test. This measure was originally validated for use in screening for COPD but comprises a set of questions routinely used by clinicians to evaluate patients for respiratory disorders.^19^ The maximum number of respiratory symptoms that could be selected by a respondent was five and the minimum number of symptoms was zero.

Cigarette status was self reported and categorized into (never smoked a cigarette, smoked a cigarette but not a current daily smoker, and daily smoker). Participants also reported number of cigarettes smoked per day, and whether or not they were past month users of alcohol and cannabis. Participants were asked what vaping device they had last used, and how long they had been vaping at the time of survey completion. Data on demographic characteristics were also collected (sex, age, education, parental status, marital status, province of residence, and race). Participants also reported whether or not they had received a diagnosis of asthma.

### Statistical Analysis

This analysis assessed the effect of e-cigarette use and frequency on the rate ratios of respiratory symptoms, assessed via a Poisson regression model. The interaction between e-cigarette puffs per day and number of cigarettes smoked per day on respiratory symptoms was also assessed. All other covariates in the model were chosen based on past research. Covariates were included in the analysis if they were deemed to be conceptually relevant as confounders. Covariates were respondent age, sex, race, education level, race, province and history of substance-use other than cigarettes or e-cigarettes.

An additional Poisson regression analysis was conducted which assessed effects of self-reported vaping device type, vaping flavour, and number of years since starting vaping on respiratory symptoms among current e-cigarette users. Analyses were performed using Stata/IC 16.1.

## Results

Descriptive statistics were calculated for all variables included in the Poisson regression model and outlined in Table 1. The average age of participants included in this analysis was 19.6 years (SD=2·7), ranging from 16 to 25 years. The majority of respondents were white (n=2270, 73.7%) and 80.6% were female (n=2456). 396 individuals reporting a diagnosis of asthma. Descriptive statistics for all analyzed variables can be seen in Table 1.

**Table 1.** Demographics for all variables included in the multivariable linear regression model.

|  | Total | Never-use of both smoking and vaping | Non-daily ever-use of either smoking or vaping | Daily vaping, non-daily/never smoking | Daily smoking, non-daily/never vaping | Daily vaping, daily smoking |
| --- | --- | --- | --- | --- | --- | --- |
|  | N=3,082 | N=825 | N=1,303 | N=820 | N=92 | N=42 |
| **Age** | 19.6 (2.7) | 18.8 (2.6) | 20.1 (2.8) | 19.6 (2.5) | 21.6 (2.6) | 19.6 (2.5) |
| **16-19** | 1,689 (54.8%) | 561 (68.0%) | 634 (48.7%) | 448 (54.6%) | 22 (23.9%) | 24 (57.1%) |
| **20-25** | 1,393 (45.2%) | 264 (32.0%) | 669 (51.3%) | 372 (45.4%) | 70 (76.1%) | 18 (42.9%) |
| **Sex** |  |  |  |  |  |  |
| **Male** | 593 (19.4%) | 116 (14.2%) | 219 (17.0%) | 227 (28.0%) | 17 (18.9%) | 14 (33.3%) |
| **Female** | 2,456 (80.6%) | 701 (85.8%) | 1,071 (83.0%) | 583 (72.0%) | 73 (81.1%) | 28 (66.7%) |
| **Race** |  |  |  |  |  |  |
| **White** | 2,270 (73.7%) | 494 (59.9%) | 983 (75.4%) | 682 (83.2%) | 78 (84.8%) | 33 (78.6%) |
| **Black** | 62 (2.0%) | 22 (2.7%) | 32 (2.5%) | 8 (1.0%) | 0 (0.0%) | 0 (0.0%) |
| **Chinese** | 201 (6.5%) | 109 (13.2%) | 72 (5.5%) | 20 (2.4%) | 0 (0.0%) | 0 (0.0%) |
| **Filipino** | 43 (1.4%) | 18 (2.2%) | 15 (1.2%) | 10 (1.2%) | 0 (0.0%) | 0 (0.0%) |
| **Indigenous** | 63 (2.0%) | 5 (0.6%) | 26 (2.0%) | 22 (2.7%) | 6 (6.5%) | 4 (9.5%) |
| **Japanese** | 4 (0.1%) | 0 (0.0%) | 3 (0.2%) | 1 (0.1%) | 0 (0.0%) | 0 (0.0%) |
| **Korean** | 19 (0.6%) | 7 (0.8%) | 7 (0.5%) | 4 (0.5%) | 1 (1.1%) | 0 (0.0%) |
| **Latin-Central-South American** | 55 (1.8%) | 12 (1.5%) | 29 (2.2%) | 11 (1.3%) | 2 (2.2%) | 1 (2.4%) |
| **Southeast Asian** | 38 (1.2%) | 21 (2.5%) | 12 (0.9%) | 5 (0.6%) | 0 (0.0%) | 0 (0.0%) |
| **South Asian** | 182 (5.9%) | 99 (12.0%) | 65 (5.0%) | 16 (2.0%) | 2 (2.2%) | 0 (0.0%) |
| **West Asian or Arab** | 56 (1.8%) | 14 (1.7%) | 30 (2.3%) | 9 (1.1%) | 1 (1.1%) | 2 (4.8%) |
| **Other** | 89 (2.9%) | 24 (2.9%) | 29 (2.2%) | 32 (3.9%) | 2 (2.2%) | 2 (4.8%) |
| **Province** |  |  |  |  |  |  |
| **Ontario** | 1,531 (49.7%) | 478 (57.9%) | 664 (51.0%) | 322 (39.3%) | 49 (53.3%) | 18 (42.9%) |
| **Alberta** | 496 (16.1%) | 104 (12.6%) | 203 (15.6%) | 168 (20.5%) | 15 (16.3%) | 6 (14.3%) |
| **British Columbia** | 483 (15.7%) | 111 (13.5%) | 210 (16.1%) | 144 (17.6%) | 10 (10.9%) | 8 (19.0%) |
| **Manitoba** | 116 (3.8%) | 25 (3.0%) | 52 (4.0%) | 32 (3.9%) | 6 (6.5%) | 1 (2.4%) |
| **New Brunswick** | 53 (1.7%) | 15 (1.8%) | 14 (1.1%) | 22 (2.7%) | 2 (2.2%) | 0 (0.0%) |
| **Newfoundland and Labrador** | 40 (1.3%) | 6 (0.7%) | 16 (1.2%) | 17 (2.1%) | 0 (0.0%) | 1 (2.4%) |
| **Nova Scotia** | 102 (3.3%) | 27 (3.3%) | 47 (3.6%) | 24 (2.9%) | 2 (2.2%) | 2 (4.8%) |
| **Prince Edward Island** | 16 (0.5%) | 2 (0.2%) | 5 (0.4%) | 9 (1.1%) | 0 (0.0%) | 0 (0.0%) |
| **Quebec** | 129 (4.2%) | 28 (3.4%) | 59 (4.5%) | 34 (4.1%) | 3 (3.3%) | 5 (11.9%) |
| **Saskatchewan** | 113 (3.7%) | 27 (3.3%) | 33 (2.5%) | 47 (5.7%) | 5 (5.4%) | 1 (2.4%) |
| **Northwest Territories** | 2 (0.1%) | 1 (0.1%) | 0 (0.0%) | 1 (0.1%) | 0 (0.0%) | 0 (0.0%) |
| **Yukon** | 1 (0.0%) | 1 (0.1%) | 0 (0.0%) | 0 (0.0%) | 0 (0.0%) | 0 (0.0%) |
| **Highest education level** |  |  |  |  |  |  |
| **Some elementary or high school** | 764 (24.8%) | 286 (34.7%) | 261 (20.0%) | 185 (22.6%) | 21 (22.8%) | 11 (26.2%) |
| **Completed High school** | 1,489 (48.3%) | 356 (43.2%) | 604 (46.4%) | 469 (57.2%) | 40 (43.5%) | 20 (47.6%) |
| **College diploma** | 268 (8.7%) | 33 (4.0%) | 133 (10.2%) | 79 (9.6%) | 18 (19.6%) | 5 (11.9%) |
| **University or post graduate degree** | 561 (18.2%) | 150 (18.2%) | 305 (23.4%) | 87 (10.6%) | 13 (14.1%) | 6 (14.3%) |
| **Marital status** |  |  |  |  |  |  |
| **Single** | 2,577 (83.6%) | 768 (93.1%) | 1,058 (81.2%) | 658 (80.2%) | 63 (68.5%) | 30 (71.4%) |
| **Married or living with a partner** | 501 (16.3%) | 55 (6.7%) | 244 (18.7%) | 162 (19.8%) | 28 (30.4%) | 12 (28.6%) |
| **Divorced/Separated/Widowed** | 4 (0.1%) | 2 (0.2%) | 1 (0.1%) | 0 (0.0%) | 1 (1.1%) | 0 (0.0%) |
| **Parental status** |  |  |  |  |  |  |
| **Yes** | 60 (1.9%) | 4 (0.5%) | 28 (2.1%) | 20 (2.4%) | 8 (8.7%) | 0 (0.0%) |
| **No** | 3,022 (98.1%) | 821 (99.5%) | 1,275 (97.9%) | 800 (97.6%) | 84 (91.3%) | 42 (100.0%) |
| **Vaping device type (last used)** |  |  |  |  |  |  |
| **Modifiable/tubular device** | 331 (15.9%) |  | 180 (15.6%) | 122 (15.2%) | 23 (25.8%) | 6 (14.3%) |
| **Pod system** | 1,426 (68.3%) |  | 753 (65.1%) | 599 (74.9%) | 47 (52.8%) | 27 (64.3%) |
| **Other<sup>1</sup>** | 331 (15.9%) |  | 224 (19.4%) | 79 (9.9%) | 19 (21.3%) | 9 (21.4%) |
| **E-liquid flavour (last used)** |  |  |  |  |  |  |
| **Fruit/Candy/Dessert/Food** | 1,351 (64.0%) |  | 737 (62.5%) | 531 (66.5%) | 54 (60.7%) | 29 (69.0%) |
| **Beverage** | 35 (1.7%) |  | 23 (1.9%) | 9 (1.1%) | 1 (1.1%) | 2 (4.8%) |
| **Mint/Menthol** | 387 (18.3%) |  | 175 (14.8%) | 192 (24.0%) | 13 (14.6%) | 7 (16.7%) |
| **Tobacco** | 59 (2.8%) |  | 23 (1.9%) | 25 (3.1%) | 8 (9.0%) | 3 (7.1%) |
| **Other** | 82 (3.9%) |  | 42 (3.6%) | 34 (4.3%) | 5 (5.6%) | 1 (2.4%) |
| **I don't know** | 196 (9.3%) |  | 180 (15.3%) | 8 (1.0%) | 8 (9.0%) | 0 (0.0%) |
| **How often do you currently vape?** |  |  |  |  |  |  |
| **Daily or almost daily** | 862 (28.0%) | 0 (0.0%) | 0 (0.0%) | 820 (100.0%) | 0 (0.0%) | 42 (100.0%) |
| **Less than daily, but at least once a week** | 178 (5.8%) | 0 (0.0%) | 154 (11.8%) | 0 (0.0%) | 24 (26.1%) | 0 (0.0%) |
| **Less than weekly, but at least once a month** | 165 (5.4%) | 0 (0.0%) | 150 (11.5%) | 0 (0.0%) | 15 (16.3%) | 0 (0.0%) |
| **Less than monthly** | 323 (10.5%) | 0 (0.0%) | 302 (23.2%) | 0 (0.0%) | 21 (22.8%) | 0 (0.0%) |
| **Not at all** | 622 (20.2%) | 0 (0.0%) | 593 (45.5%) | 0 (0.0%) | 29 (31.5%) | 0 (0.0%) |
| **I have never vaped** | 932 (30.2%) | 825 (100.0%) | 104 (8.0%) | 0 (0.0%) | 3 (3.3%) | 0 (0.0%) |
| **Cigarettes smoked daily** | 4.4 (7.2) |  | 1.9 (3.0) | 3.2 (4.7) | 9.8 (11.8) | 5.7 (5.9) |
| **Daily alcohol/cannabis use** | 878 (28.5%) | 30 (3.6%) | 348 (26.7%) | 420 (51.2%) | 51 (55.4%) | 29 (69.0%) |
| **Asthma diagnosis** | 396 (12.8%) | 107 (13.0%) | 154 (11.8%) | 106 (12.9%) | 19 (20.7%) | 10 (23.8%) |

<sup>1</sup>Other devices: disposable or rechargeable cigarette-like vaping devices, pen-like devices, and other.

Half of respondents reported either never vaping or not currently vaping at all (n=1554, 50.4%), 10.5% reported less than monthly vaping (n=323), 5.4% reported vaping monthly (n=165), 5.8% vaped weekly (n=178), and 28.0% reported vaping daily (n=862). The average number of cigarettes smoked per day among ever-smokers in the sample was 4.4 (SD=7·2), while the daily average among exclusive current daily smokers was 9.8 (SD=11.8) and the daily average among current daily dual users was 5.7 (SD=5.9) cigarettes per day. Never smokers who vaped had lower numbers of puffs per day, number of days vaped per month, and number of years vaped than ever smokers and daily smokers (Table 2).

**Table 2.** Mean number of puffs per day vaped, number of days per month vaped, number of years vaped, by smoking status (never, ever, and daily) among ever vapers (n=2150)

|  | Never Smokers<br>(n=840) |  | Ever Smokers (except daily)<br>(n=1118) |  | Daily Smoker<br>(n=192) |  |
| --- | --- | --- | --- | --- | --- | --- |
|  | Mean | SD | Mean | SD | Mean | SD |
| Number of puffs per day | 3.2 | 0.2 | 5.9 | 0.2 | 7.8 | 0.7 |
| Number of days vaped | 6.4 | 0.4 | 12.6 | 0.4 | 13.6 | 1 |
| Number of years vaped | 1.7 | 0.1 | 2.4 | 0.1 | 2.2 | 0.2 |

Table 3 outlines the percentage distribution of respiratory symptoms by smoking and vaping status. Half of respondents reported zero respiratory symptoms (51.5%). Of never-users, non-daily and daily e-cigarette users, the highest proportion of respondents reported zero symptoms (65.8%, 56.8%, and 34.2%, respectively), compared to the highest proportion of daily smokers who reported experiencing one respiratory symptom (23.2%), and the highest proportion of dual-users who reported experiencing two respiratory symptoms (25.0%). While for three of the five symptoms the majority of participants did not report experiencing the symptom, regardless of smoking or vaping status, the majority of daily smokers, regardless of vaping status, did report coughing regularly in the last four months (65.6% of daily smokers, non-vapers; 57.1% of daily smokers, daily vapers), and the majority of daily smokers, non-vapers reported coughing up phlegm regularly (52.2%). Additionally, daily smokers, daily vapers, and daily dual-users were all more likely to report experiencing any of the five symptoms compared to non-daily or never-users of either cigarettes or e-cigarettes. Daily smokers were also more likely to report experiencing any symptoms compared to daily vapers. Dual users had lower rates of reporting of all symptoms compared to daily smoking alone, especially when looking at wheezing (35.8% of dual-users, compared to 46.1% of daily smokers) (p<0.05 for cold frequency and duration, p<0.001 for all other respiratory symptoms and total number of symptoms).

**Table 3:** Descriptive statistics of percentage of participants with respiratory symptoms by smoking and vaping status.

|  | Total | Never-use of both smoking and vaping | Non-daily ever-use of either smoking or vaping | Daily vaping, non-daily/never smoking | Daily smoking, non-daily/never vaping | Daily vaping, daily smoking | p-value |
| --- | --- | --- | --- | --- | --- | --- | --- |
|  | N=3,082 | N=825 | N=1,303 | N=820 | N=92 | N=42 |  |
| **Number of respiratory symptoms N= 2,973** |  |  |  |  |  |  | <0.001 |
| **0** | 1,531 (51.5%) | 534 (65.8%) | 712 (56.8%) | 266 (34.2%) | 12 (13.3%) | 7 (17.5%) |  |
| **1** | 683 (23.0%) | 168 (20.7%) | 277 (22.1%) | 209 (26.9%) | 21 (23.3%) | 8 (20.0%) |  |
| **2** | 425 (14.3%) | 76 (9.4%) | 166 (13.2%) | 156 (20.1%) | 17 (18.9%) | 10 (25.0%) |  |
| **3** | 186 (6.3%) | 21 (2.6%) | 66 (5.3%) | 77 (9.9%) | 17 (18.9%) | 5 (12.5%) |  |
| **4** | 111 (3.7%) | 11 (1.4%) | 23 (1.8%) | 53 (6.8%) | 16 (17.8%) | 8 (20.0%) |  |
| **5** | 37 (1.2%) | 1 (0.1%) | 10 (0.8%) | 17 (2.2%) | 7 (7.8%) | 2 (5.0%) |  |
| **Over the past 4 months, did you cough regularly? N=2,999** |  |  |  |  |  |  | <0.001 |
| **Yes** | 686 (22.9%) | 81 (9.9%) | 207 (16.4%) | 315 (40.1%) | 59 (65.6%) | 24 (57.1%) |  |
| **Over the past 4 months, did you cough up phlegm regularly? N=2,996** |  |  |  |  |  |  | <0.001 |
| **Yes** | 509 (17.0%) | 67 (8.2%) | 170 (13.4%) | 206 (26.3%) | 47 (52.2%) | 19 (45.2%) |  |
| **Over the past 4 months, did even simple chores make you short of breath? N=2,992** |  |  |  |  |  |  | <0.001 |
| **Yes** | 629 (21.0%) | 94 (11.5%) | 231 (18.3%) | 242 (30.9%) | 41 (45.6%) | 21 (51.2%) |  |
| **Over the past 4 months, did you wheeze when you exert yourself? N=2,995** |  |  |  |  |  |  | <0.001 |
| **Yes** | 673 (22.5%) | 135 (16.5%) | 254 (20.1%) | 223 (28.4%) | 43 (47.8%) | 18 (43.9%) |  |
| **Over the past 4 months, did you get many colds and do your colds usually last longer than your friends' colds? N=2,999** |  |  |  |  |  |  | <0.05 |
| **Yes** | 247 (8.2%) | 61 (7.5%) | 97 (7.6%) | 70 (8.9%) | 15 (16.7%) | 4 (9.8%) |  |

Each additional puff year increased the rate ratio of respiratory symptoms by a factor of 11.36 (95%CI: 4.61-28.00; p<0.001) for never smokers and ever smokers (2.79; 95% CI: 1.69, 4.61), but among current daily smokers higher pack equivalent years were not associated with more respiratory symptoms (0.83; 95% CI: 0.23., 3.11) (Table 4). Test of interactions were statistically significant (p<0.001) (See supplemental files).

**Table 4.** Poisson regression summary table of rate ratios for effects of vaping and smoking frequencies on respiratory symptoms stratified by smoking status)

|  | Non Smokers | Ever Smokers | Daily Smokers |
| --- | --- | --- | --- |
| **Respiratory symptoms** |  |  |  |
| Pack equivalent years | 11.36***<br>[4.61,28.00] | 2.79***<br>[1.69,4.61] | 0.84<br>[0.23,3.11] |
| *N* | 1445 | 1059 | 156 |

<sup>2</sup>Exponentiated coefficients adjusted for respondent age, sex, education, marital status, parental status, registration date, other daily drug use, province, and race, those who reported a diagnosis of asthma are excluded. Full tables with covariates available in Supplementary Materials (Supplementary Table 1).
\* p<0.05, \*\* p<0.01, \*\*\* p<0.001

Figure 1 depicts the modelled relationship between pack equivalent years and number of respiratory symptoms, by cigarette smoking status. Never smokers with no vaping history have fewer respiratory symptoms than those who have smoked with average number of symptoms of respiratory symptoms approaches the level seen in daily smokers after 0.5 puff year. Daily smokers do not demonstrate increasing symptoms with greater pack equivalent years.

**Figure 1.**
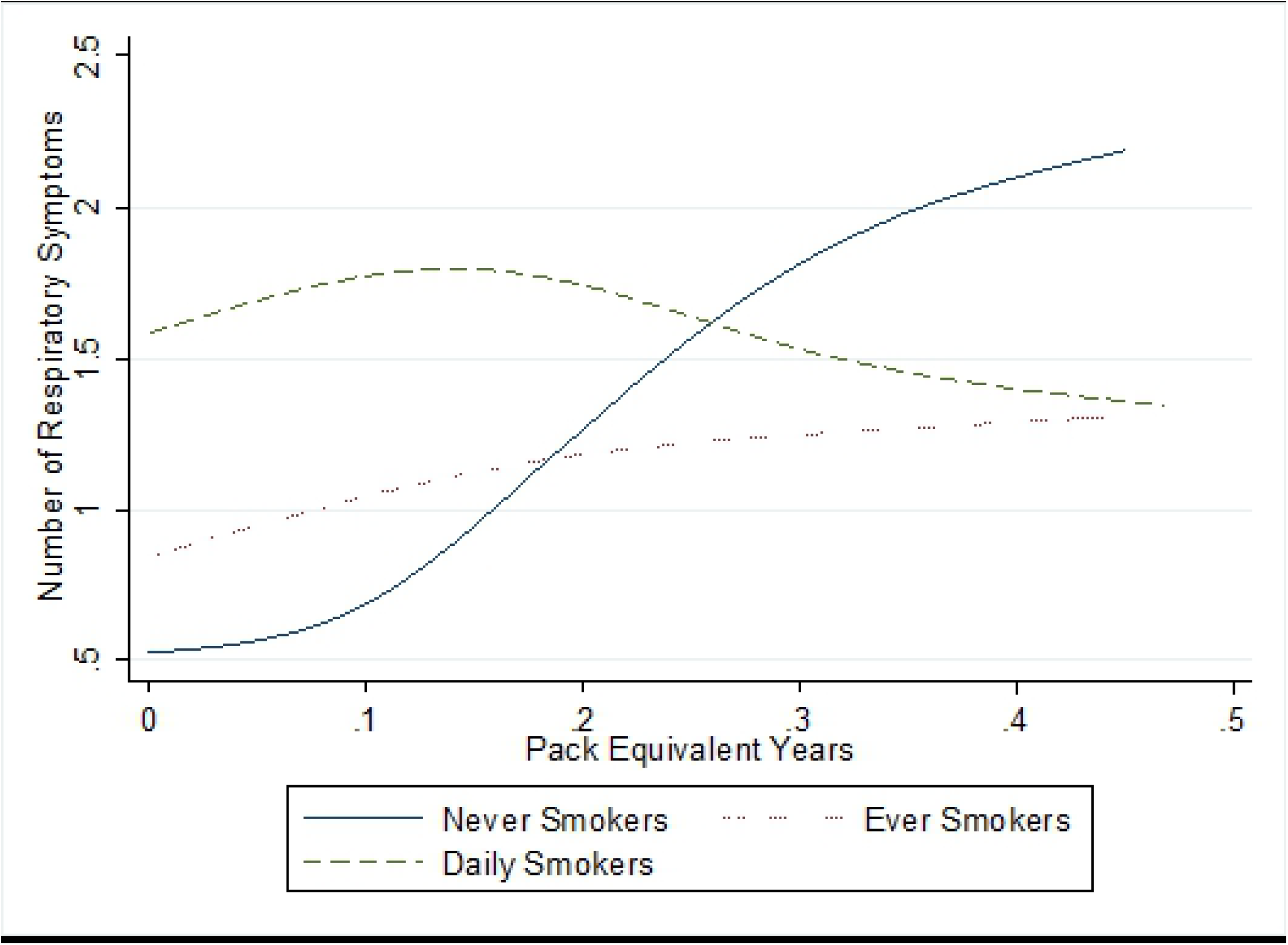
Modelled number of respiratory symptom by e-cigarette pack equivalent years by smoking status among Canadian youth (16-25) (n=2660)

Among current vapers, those using pod-style devices were more likely to have more respiratory symptoms (1.25; 95% CI: 1.08, 1.45) after adjusting for dose (See supplemental files).

## Discussion

Our analysis found that use of e-cigarettes was associated with increased rate of respiratory symptoms, and that the greater the frequency of vaping the higher the number of symptoms.

Unsurprisingly, higher amounts of cigarette smoking were also strongly associated with more respiratory symptoms. This population of youth and young adults reflects a lighter smoking sample than would be found among all adults. Exclusive daily smokers reported smoking an average of 9.8 cigarettes per day, while dual users reported smoking an average of 5.7 cigarettes per day, which is notably lower than previous research that showed an average daily cigarette consumption among Canadian daily smokers of 13.7 in 2017.^20^ Despite this level of smoking, elevated respiratory symptoms were higher in smokers than non-smokers indicating the dangerousness of cigarettes. Similarly, this population of youth and young adults have limited number of years of experience vaping, but despite this, increased frequency and length of time vaping was associated with increased numbers of respiratory symptoms that can, with sufficient dose, approach the short term respiratory harms associated with smoking cigarettes.

With regards to dual use, however, we observed that those individuals who vaped daily and smoked cigarettes daily did not demonstrate additional risk from vaping. That is, dual users reported notably high levels of respiratory symptoms compared to non-users of both cigarettes and e-cigarettes, though an increase in e-cigarette puffs resulted in a marginal decrease in rate of respiratory symptoms. These results are congruent with existing studies that reported health benefits of dual use compared to exclusive cigarette smoking.^12,13,15,17^ However, due to the high baseline of respiratory symptoms among cigarette smokers, it is difficult to ascertain the true benefit of dual-use without longitudinally monitoring changes in smoking behaviours of exclusive smokers that become dual users, to determine the mechanism behind this decrease in symptoms.

This study also builds on these results via the inclusion of a vaping-only group, as well as via the inclusion of youth participants, which allows for more specified results and interpretations.

Though the specification of age may limit generalizability to the overall population, it also allows for increased accuracy and reliability in constructing prevention and treatment plans for youth. Additional studies that monitor exact number of cigarettes smoked and e-cigarette puffs among single users versus dual users are needed to confirm this.

Those who vaped pod like devices reported higher levels of symptoms after controlling for puff year. This finding is consistent with previous results wherein adolescents reported experiencing worse respiratory symptoms when using specific vape brands and products, particularly JUUL, a pod device with a nicotine salt liquid.^21^ As was expected, subjects with a longer history of vaping experienced more respiratory symptoms than newer vapers, as they have likely inhaled a larger number of total puffs.

### Limitations

While this analysis provided insight into the association between the dual use of vaping and smoking on respiratory health symptoms, there are limitations that should be noted. First, there are substantial limitations to the generalizability of this survey due to the purposive online sampling. The high participation of females suggests that the results are more robust for young females. While the anonymous, online nature of the survey may encourage honesty, there is also a risk of recall bias, as participants may incorrectly recall their use behaviours or respiratory symptoms, either unintentionally or intentionally, especially if they are advocates for smoking or vaping. Additional studies that include physical assessments of subjects’ respiratory health and biomarkers of smoking/vaping are necessary to achieve more accurate and reliable results. The cross-sectional nature of these data also presents some limitations with regards to the conclusions that can be drawn from our analysis. Because survey respondents were asked about their use of cigarettes and e-cigarettes simultaneously and retrospectively, it is unclear how patterns of use changed over the lifecourse and how these reported symptoms may or may not lead to medical diagnosis of disease long term.

## Conclusion

This study documented a potential correlation between vaping and an increase in respiratory symptoms. A secondary finding was an interaction between dual use of cigarettes and e-cigarettes and a decreased risk of respiratory symptoms compared to cigarette smoking alone, which exhibited a higher baseline of symptoms. Further longitudinal research using physical health assessments and biomarkers is necessary to explore this association further.

## Data Availability

De-identified participant data can be made available upon request to researchers with permission from an academic ethics review board.

## Acknowledgements

Dr. Eissenberg is a paid consultant in litigation against the tobacco industry and also the electronic cigarette industry and is named on one patent for a device that measures the puffing behavior of electronic cigarette users and on another patent for a smartphone app that determines electronic cigarette device and liquid characteristics. Other authors have no disclosures to report. The CSTP is supported by grant number U54DA036105 from the National Institute on Drug Abuse of the National Institutes of Health and the Center for Tobacco Products of the U.S. Food and Drug Administration. The content of this message is solely the responsibility of the author and does not necessarily represent the views of the NIH or the FDA The cohort study was funded by CIHR. The funding sources had no role in the study design, collection, analysis, and interpretation of data, writing the report and the decision to submit the report for publication.

